# Preconception Care for Women with Medicaid: Self-report vs. Claims-based Utilization Measures

**DOI:** 10.1101/2022.07.04.22277141

**Authors:** Debra B. Stulberg, L. Philip Schumm, Kellie Schueler, Mihai Giurcanu, Monica Peek

## Abstract

**Background:** Preconception care may improve perinatal outcomes and reduce disparities, but there is no standard population measure of preconception care utilization (PCU).

**Objective:** We compared claims-based PCU from Medicaid Analytic Extract (MAX) data to self-report in the Pregnancy Risk Assessment and Monitoring System (PRAMS) survey.

**Methods:** Among Medicaid-enrolled women ages 15-45 with births during 2012, we identified preconception services in MAX using 55 ICD9 codes published by Health and Human Services. We estimated the proportion reporting preconception care from 26 PRAMS states and compared this to the states’ proportion who received services in MAX. We fit mixed-effects logistic regression models of the probability of PCU on demographic factors (age, race/ethnicity) and diagnoses (depression, diabetes, or hypertension), separately for each dataset. Finally, we computed the population proportions receiving care by state (MAX) and the empirical Bayes means of the state-level effects (MAX and PRAMS).

**Results:** Among 652,929 deliveries in MAX from the included states, 28.1% received at least one preconception service. In PRAMS, 23.6% (95% CI [22.1, 25.3]) of Medicaid-covered respondents reported preconception care. In both datasets, PCU rates were higher for Black non-Hispanic vs. White non-Hispanic women (PRAMS OR 2.05 [1.60, 1.62]; MAX OR 1.51 [1.49, 1.54]) and for those with diabetes (PRAMS OR 1.82 [1.16, 2.85]; MAX OR 1.34 [1.29, 1.40]) or hypertension (OR 1.85 [1.41, 2.44]; MAX OR 1.22 [1.18, 1.27]). In PRAMS, Asian (OR 3.37 [2.28, 4.98]) and Hispanic women (OR 2.07 [1.5, 2.80]) were more likely to report PCU than White non-Hispanic women, but in MAX they were less likely to receive services. The correlation between the PRAMS state-specific effects and those from MAX was 0.31 (p = 0.124).

**Conclusions:** Claims-based estimates of PCU are moderately concordant with self-reported rates at the state level; however, rates measured through Medicaid claims vs. self-report diverge in some groups.

**Synopsis:** *Study Question:* How do Medicaid claims-based measures of preconception care utilization compare to self-reported receipt of preconception counseling among Medicaid-covered respondents in the Pregnancy Risk Assessment and Monitoring System (PRAMS) survey?

*What’s already known:* PRAMS provides population-level estimates of preconception care utilization, while claims-based measures quantify specific services received.

*What this study adds:* Claims-based preconception care utilization among the Medicaid population varies by race (Black/White) and diagnosis of diabetes or hypertension in similar patterns as self-report in PRAMS, but ethnicity (Hispanic/non-Hispanic) and depression demonstrate divergent patterns between the two data sources. State-level variation in preconception care utilization is greater in claims data. Both data sources can be used by researchers with an understanding of their methodological benefits and limitations.

## Background

Preconception care, defined as preventative healthcare a patient receives before pregnancy to address pregnancy-related risk factors, has been hailed as a promising strategy to improve maternal and infant outcomes and to reduce health disparities.^1-4^ The Centers for Disease Control and Prevention (CDC) recommends that all women of childbearing age receive preconception care.^3^ The theoretical basis for preconception care is that many underlying causes of adverse pregnancy outcomes, such as maternal chronic diseases, are best addressed using a preventative approach before pregnancy.^5^ While high quality prenatal and intrapartum care are important, they may occur too late to mitigate many risk factors and to prevent maternal and infant morbidity and mortality. The Integrated Perinatal Health Framework emphasizes the role of multiple determinants on pregnancy outcomes.^6^

Specific preconception interventions are supported by randomized controlled trials (RCTs) demonstrating, for example, that preconception glycemic control for women with diabetes improves infant outcomes, and that preconception counseling improves maternal behaviors before pregnancy such as reducing alcohol consumption and increasing folic acid intake, which improves infant outcomes.^7-10^ Furthermore, preconception care may reduce racial and ethnic disparities in adverse pregnancy outcomes, since multiple preconception risk factors are disproportionately prevalent among women of color.^11^

Self-reported receipt of preconception care – both general counseling and specific preconception health services – is assessed on several population surveys, including the CDC’s Pregnancy Risk Assessment and Monitoring Systems (PRAMS) and the Behavioral Risk Factor Surveillance System (BRFSS).^12,13^ However, for population surveillance, reliable measures of preconception care utilization (PCU) drawn from administrative sources that do not rely on self-report would be beneficial.

## Methods

### Data

We conducted a retrospective secondary analysis of Medicaid Analytic Extract (MAX) data files from the Centers for Medicare and Medicaid Services (CMS) from 2010-2012 under an approved Data Use Agreement. These data files include person-level information on Medicaid enrollees and encounter-level information for Medicaid claims from all sources of care, including inpatient, outpatient, physician services, radiology, clinic visits, and pharmacies. The University of Chicago’s Institutional Review Board approved this study.

We reviewed claims from all female beneficiaries, aged 15-45, enrolled in Medicaid from all available states (45) and Washington DC who experienced a delivery in 2012. Deliveries were identified using the following International Classification of Diseases-9^th^ revision (ICD9) diagnosis codes: V27.xx with or without 650 for normal deliveries; and V27.xx with 644.2, 644.4, 765.0 or 765.1 for preterm births. For women with more than one delivery in calendar year 2012, only information from the first delivery was used.

For each index 2012 birth, the corresponding date of conception was estimated using a modified version of the approach described by Palmsten et al.^14^ The date of conception was calculated to be 255 days before a full-term birth, 230 days before a premature birth. We identified preconception care in the MAX Other Therapies (OT) files using a list of 55 International Classification of Diseases-9^th^ Revision (ICD9) codes published by the United States Department of Health and Human Services’ Office of Population Affairs under its Quality Family Planning program.^15^ These are classified in 7 domains of services (Table 1): contraceptive services, pregnancy testing and counseling, achieving pregnancy, basic infertility services, preconception health services, sexual transmitted diseases services, and related preventative health services. Examples of these service categories include v25.01 (counseling on oral contraceptive prescription) under contraceptive services, and v70.0 (general medical exam) under related preventive health services. If a woman had an encounter in the year prior to conception that included any diagnosis code from any of these 7 domains, we classified her as having received preconception care. We also computed separate indicators of having received care within each of the 7 preconception care domains.

**Table 1.** ICD9 codes included in each preconception care domain

| Preconception care domain | ICD9 Codes |
| --- | --- |
| Contraceptive services | V25.01, V25.02, V25.03, V25.04, V25.09, V25.11, V25.12, V25.13, V25.2, V25.40, V25.41, V25.42, V25.43, V25.49, V25.5, V25.8, V25.9 |
| Pregnancy testing and counseling | V72.40, V72.41, V72.42 |
| Achieving pregnancy | V26.41 |
| Basic infertility services | 606.9, 628.0, 628.1, 628.2, 628.3, 628.4, 628.9, V26.21 |
| Preconception health services | V15.82, V26.49, V65.3, V65.41, V65.42, V77.1, V77.8, V79.0, V79.1, V81.1 |
| STD services | V01.6, V02.8, V12.09, V65.40, V65.44, V65.45, V65.5, V69.2, V73.81, V73.88, V73.89, V74.5 |
| Related preventative health services | V70.0, V72.31, V76.19, V76.2 |

We compared the claims-based measure to patient reported receipt of preconception counseling among Medicaid-covered PRAMS respondents who experienced a delivery in 2012. We received data under an approved Data Sharing Agreement with the CDC. We used data from PRAMS Phase 7 Core Questionnaire to identify women who received Medicaid prior to pregnancy (Question 8: “During the month before you got pregnant with your new baby, what kind of health insurance did you have?”); of these, we identified those who reported receiving preconception health counseling (Question 10: “Before you got pregnant with your new baby, did a doctor, nurse, or other health care worker talk to you about how to improve your health before pregnancy?”). The data from 26 states that administered the question about preconception counseling in 2012 were used in the data analysis. Survey weights adjusting for differences in the probability of selection and differential non-response were provided with the data, as was information about the sampling strata; these were used in the analysis to obtain unbiased estimates and design-based standard errors.

Covariates hypothesized to be associated with the likelihood of receiving preconception care and available in both datasets included the following: (1) age group (≤17, 18–19, 20–24, 25–29, 30–34, 35– 39, 40+); (2) race/ethnicity (White non-Hispanic, Black non-Hispanic, Hispanic, Asian/Pacific Islander, and Other); and (3) presence of chronic conditions including diabetes, hypertension and depression (obtained from the chronic conditions file in MAX).

### Statistical Analysis

We estimated the proportion receiving preconception care from PRAMS together with an approximate 95% CI, and compared this to the population proportion who received preconception care for those same 26 states from MAX. As noted above, design-based variance estimates computed using the linearization method^16^ were used in the analysis of PRAMS throughout. We also computed the population proportions receiving care within each of the 7 care domains using MAX. Finally, we compared the survey weighted estimates of the distributions of categorical variables from PRAMS to the corresponding population values from MAX.

We fit mixed-effects logistic regression models ^17^of the probability of receiving preconception care on the demographic and clinical covariates, separately for each dataset (as before, analyses of MAX are restricted to the 26 states available in PRAMS). Random effects (intercepts) were included at the state level to accommodate additional variability between states after adjusting for the covariates. Since the standard mixed model assumes that the random, state-level effects are uncorrelated with the covariates, we included the state-level means of each covariate in the model to address the problems that can arise if this assumption is violated.^18^ Models were fit using maximum likelihood with mean– variance adaptive Gauss–Hermite quadrature.^19^ Estimated coefficients are presented as odds ratios, together with approximate 95% confidence intervals. A test of the null hypothesis that the true variance of the random effect is zero was performed using the asymptotic null distribution derived by Self and Liang (1987).^20,21^ Marginal probabilities^22^ of receiving preconception care were computed and plotted by age group for each dataset to facilitate interpretation of differences by age. Additional models excluding the random effects and including data for all 45 states and DC (MAX only) were also fit for comparison, and are included as Supplementary Materials.

To explore geographic variability in the prevalence of preconception care, we computed and plotted the population proportions (MAX) receiving care by state. We also computed the empirical Bayes means of the state-level effects and plotted these (1) on a US map, (2) against the unadjusted population proportions; and (3) for MAX against those for PRAMS.

All analyses were performed using Stata Release 17.0.^23^

## Results

Using the MAX data we identified 1,452,034 women who delivered in 2012, of whom 23.1% received at least one health care service from 7 domains of care within the year prior to conception. The percentage receiving preconception care varied across states (Figure 2a). Excluding those not living in one of the 26 PRAMS states reduced the number of women to 652,929, 28.1% of whom received preconception care (Table 2). By comparison, among 6,960 PRAMS respondents we estimated that only 23.6% (95% CI 22.1, 25.3) of Medicaid-eligible women delivering in 2012 reported receiving preconception care (this difference was concentrated below age 25 as described below). Estimated distributions for age and race/ethnicity from PRAMS were similar to those observed in MAX, though PRAMS slightly overestimated the proportion less than 20 years old and the proportion Black/African American while slightly underestimating the proportion non-Hispanic White. The prevalence of all three chronic conditions based on self-report in PRAMS was higher than those based on the chronic conditions file in MAX.

**Table 2.** Survey weighted distributions of demographic and clinical characteristics in PRAMS compared to MAX, 2012 (percentages and 95% CI)

|  | PRAMS (n=6,960)<sup>a</sup> | MAX (n=652,929)<sup>b</sup> |
| --- | --- | --- |
| Age |  |  |
| ≤17 | 5.3 (4.6, 6.1) | 3.6 |
| 18–19 | 11.8 (10.6, 13.1) | 9.1 |
| 20–24 | 32.5 (30.7, 34.4) | 35.0 |
| 25–29 | 27.2 (25.5, 28.9) | 27.9 |
| 30–34 | 15.6 (14.3, 17.0) | 16.1 |
| 35–39 | 5.9 (5.1, 6.8) | 6.6 |
| 40+ | 1.8 (1.3, 2.3) | 1.7 |
| Race/ethnicity |  |  |
| White non-Hispanic | 45.6 (44.0, 47.2) | 50.4 |
| Black non-Hispanic | 28.1 (26.6, 29.7) | 24.9 |
| Hispanic | 16.2 (15.1, 17.3) | 17.3 |
| Asian/Pacific Islander | 5.2 (4.6, 5.9) | 4.5 |
| Other | 4.9 (4.3, 5.5) | 3.0 |
| Chronic Conditions |  |  |
| Diabetes | 4.0 (3.3, 4.8) | 1.6 |
| Hypertension | 7.2 (6.3, 8.3) | 2.2 |
| Depression | 15.6 (14.3, 17.0) | 7.2 |
| Preconception care | 23.6 (22.1, 25.3) | 28.1<sup>c</sup> |
| Contraceptive services |  | 13.2 |
| Related preventative health services |  | 15.4 |

<sup>a</sup> Includes only those who were Medicaid-eligible; estimates use the survey weights provided with the dataset.
<sup>b</sup> Includes only those states for which PRAMS data were available.
<sup>c</sup> Includes all individuals with at least one claim during the year before conception in the following 7 domains: contraceptive services, pregnancy testing and counseling, achieving pregnancy, basic infertility services, preconception health services, STD services, and related preventative health services.

Table 3 shows results from mixed-effects logistic regression models fit separately to both datasets. For both PRAMS and MAX, the proportion receiving preconception care declined with age after ages 25–29 and the marginal proportions for both datasets were similar, whereas at younger ages the estimated proportions were approximately 5–10 percentage points less for PRAMS (Figure 1a). For PRAMS both Black and Hispanic respondents were substantially more likely to report having receiving preconception care than White respondents, with odds ratios of 2.05 (95% CI 1.60, 2.62) and 2.07 (95% CI 1.53, 2.80), respectively. The same was true for respondents describing themselves as Asian/Pacific Islanders or “Other.” By contrast, while Black non-Hispanic women in MAX were also more likely than White non-Hispanic women to have received preconception care with an odds ratio of 1.51 (95% CI 1.49, 1.54), Hispanic women and Asian/Pacific Islander women were actually *less likely* to have received preconception care with odds ratios of 0.74 (95% CI 0.73, 0.75) and 0.65 (95% CI 0.63, 0.67), respectively (Figure 1b). Women with diabetes and hypertension were more likely to have received preconception care in both datasets though the differences were smaller for MAX, whereas women with depression were more likely to have received preconception care in MAX only (no difference was observed for PRAMS). Results from logistic regressions excluding the random effects were similar (Supplemental Table 1).

**Table 3.**
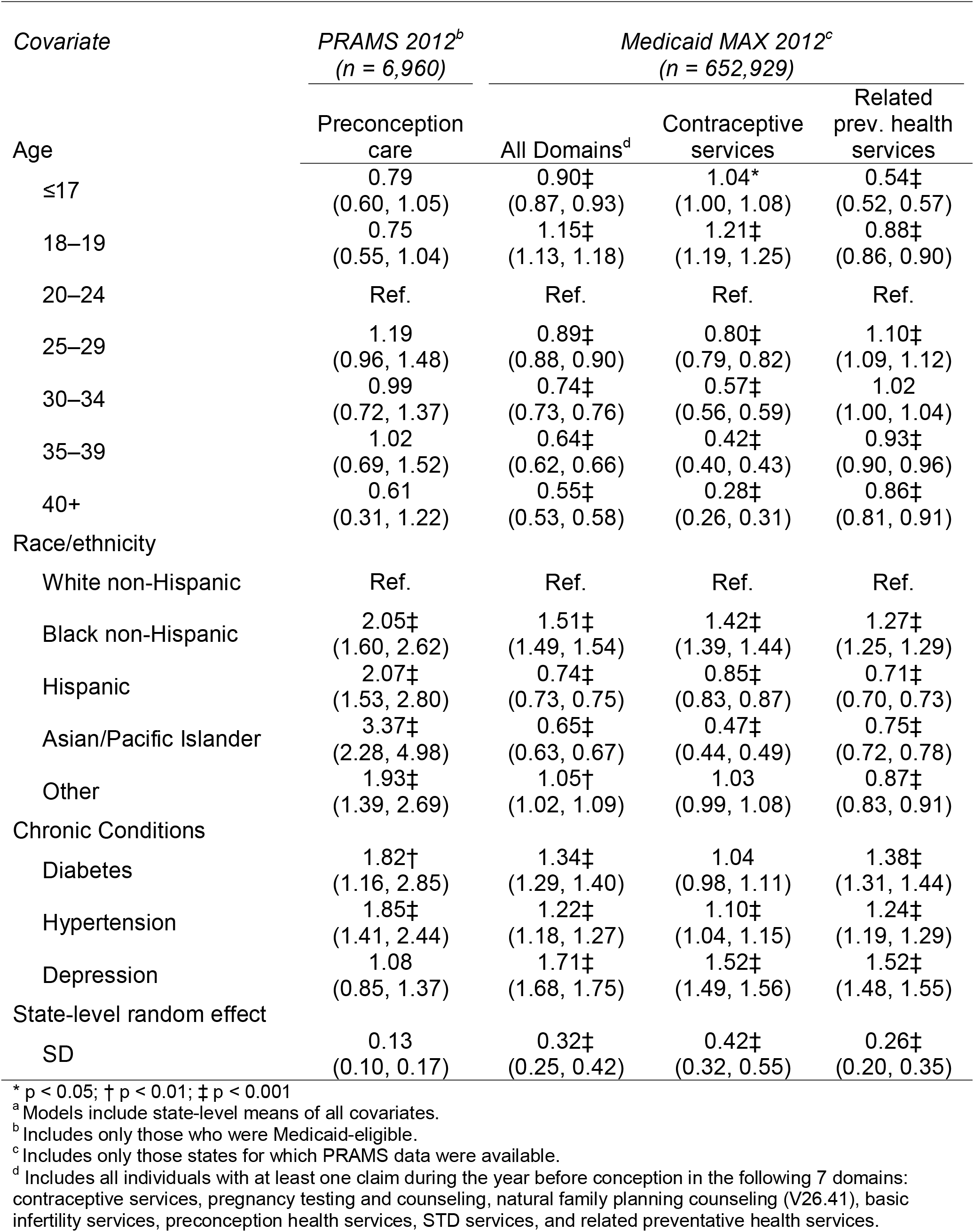
Mixed-effects logistic regressions of preconception care measures from 2012 PRAMS and Medicaid MAX on age, race/ethnicity and chronic conditions, estimated using data from 26 states available in PRAMS (odds ratios and 95% CI)^a^

**Figure 1.**
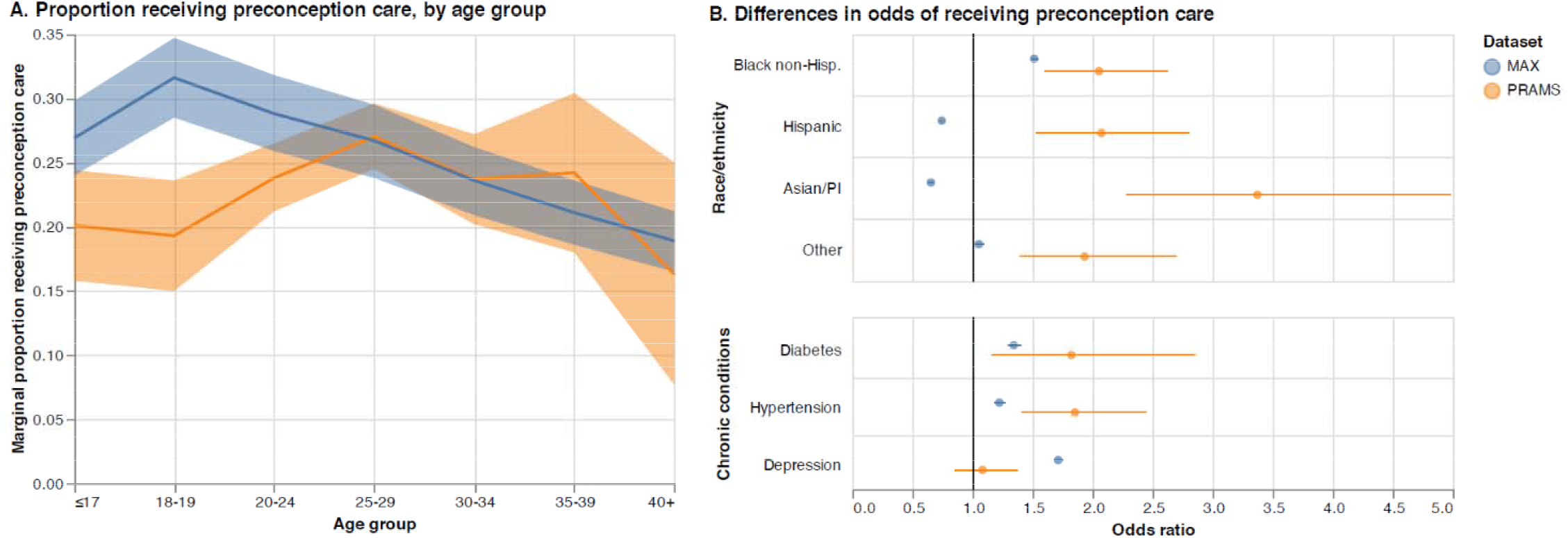
Results from mixed-effects models, Medicaid MAX and PRAMS 2012

Roughly similar patterns were also observed in MAX for the likelihood of having received contraceptive services and related preventative health services, both domains of preconception care associated with decreased risk of severe maternal morbidity.^24^ Notable differences included somewhat different patterns with age, and negligible or smaller differences in the likelihood of having received contraceptive services for those with diabetes or hypertension versus those without, with odds ratios of 1.04 (95% CI 0.98, 1.11) and 1.10 (95% CI 1.04, 1.15), respectively.

In MAX, the standard deviation of the state-level random effects in the model for all preconception care domains was 0.32 (95% CI 0.25, 0.42), corresponding to only 3% of the total variance after adjusting for the covariates. The standard deviation was slightly higher when fit to the data for all 45 MAX states plus DC at 0.42 (95% CI 0.34, 0.52) (Supplemental Table 2). Estimates of the state-specific random effects are plotted in Figure 2b; the correlation between these and the state-specific prevalence of care was 0.67 (Figure 2c). The standard deviation of the random effects for PRAMS was smaller (0.13), and a test of the null hypothesis that the state level variance component is zero was not significant (p = 0.28). Among the 26 PRAMS states, the correlation between the PRAMS state-specific effects and those from MAX was 0.31 (p = 0.124) (Figure 2d).

**Figure 2.**
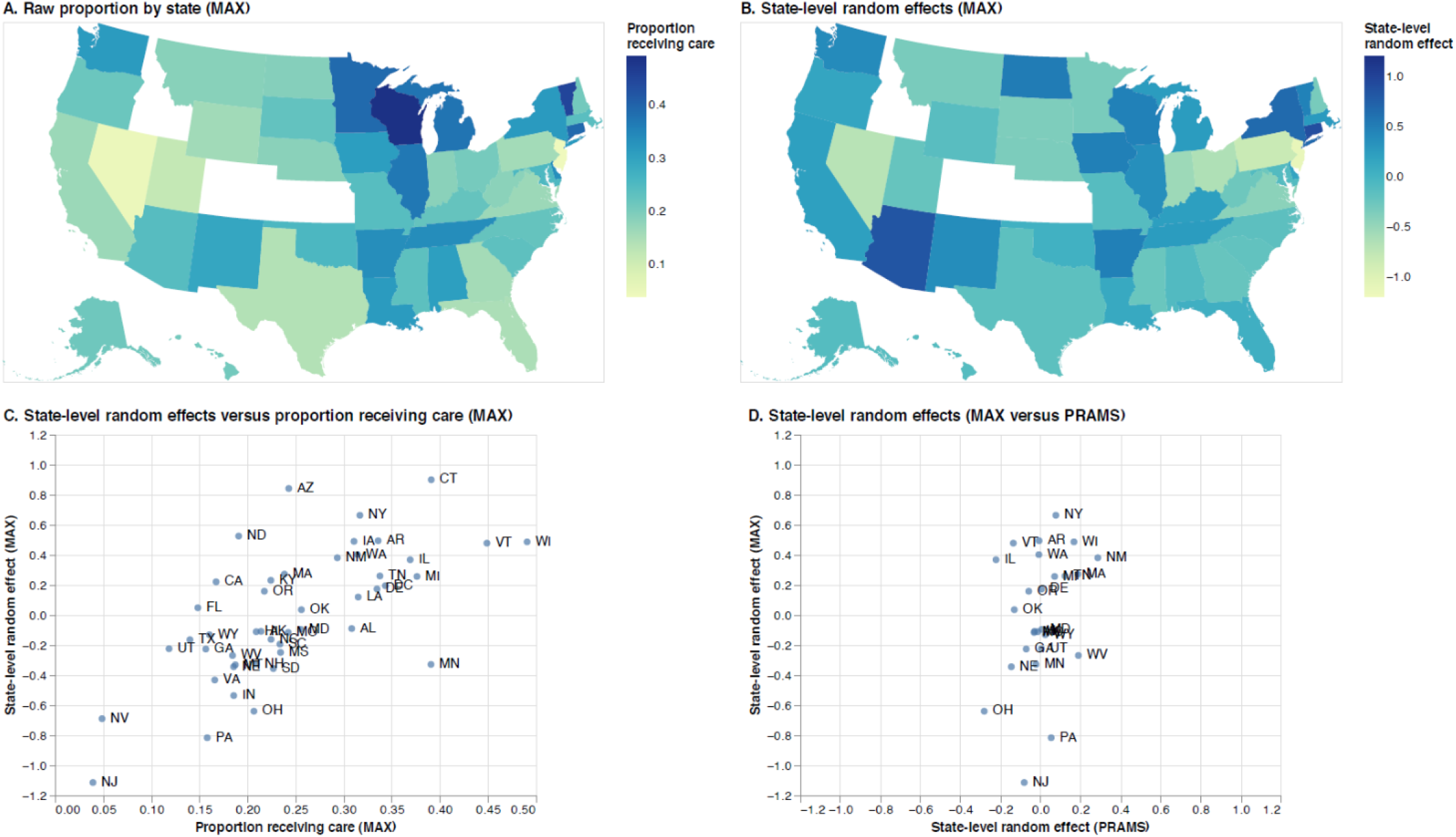
Geographic variation in prevalence or preconception care, Medicaid MAX and PRAMS 2012

### Comment

In this analysis, we compared a Medicaid claims-based measure of preconception care utilization to women’s self-report from the PRAMS survey. Overall, we found similar rates of PCU in the two datasets and similar associations between PCU and both race (PCU was higher among Black than among White women) and the presence of chronic conditions (PCU was more common among those with diabetes or hypertension). At the same time, we found that women under 25 and those with depression were less likely to report PCU than was reflected in the claims, while Hispanic women were substantially more likely to report PCU. These differences likely reflect differences between subgroups of women in the way that the single item question in PRAMS is understood, as well as differences in the types of and manner in which PCU is delivered and billed by providers. For example, women from different racial/ethnic backgrounds may have different sociocultural understandings of health and disease that affect their interactions with a provider and in turn affect how they report on those interactions. In addition, certain interactions a provider may have with a patient may correspond to billing codes that are underutilized or may have no codes at all, and these types of interactions may occur more commonly among certain subgroups of women than others. Finally, it is possible that some of the specific services delivered as part of preconception care are simply not recognized as such (or remembered) by the individual receiving them. For example, women trying to conceive may be more attuned to such services than others.

Limitations to PRAMS survey measures include lack of specificity about services received, lack of information about reproductive history, and narrowly including only women with a recent live birth.^25^ In addition, PRAMS does not assess rare but serious maternal complications such as severe maternal morbidity,^28^ which would be difficult to identify through self-report but can be quantified and assessed for association with preconception services using Medicaid claims.^24^ Furthermore, PRAMS asks postpartum women to recall if a healthcare provider counseled them about their health prior to pregnancy, which may be subject to recall bias. Because of these inherent limitations in using surveys for tracking preconception care, it is encouraging to see claims-based measures offer a promising alternative for population-wide surveillance. Nonetheless, claims data also have important limitations, such as the fact that while diagnosis and procedures codes capture an array of billed services, they do not capture the content or quality of counseling during encounters. Furthermore, variation in provider billing practices and state reporting may generate additional variation that may have little or no relation to subsequent outcomes. This may account for the additional state-level variability observed in MAX.

The divergent patterns in PCU observed here may also be due in part to features of the Medicaid program and the way in which data apart from those on PCU are collected. For example, Medicaid churn—people entering and leaving Medicaid based on changes in income or other life circumstances—is more common among Hispanic women^29^ which could cause under-counting of preconception care services received by this group. In addition, the quality of race and ethnicity data in Medicaid has been found to vary across states.^30^

PRAMS provides an important basis for obtaining population estimates of preconception care utilization among women who have given birth.^25-27^ Researchers have used PRAMS to compare PCU between racial/ethnic groups and to track improvements in women’s health and healthcare following the Affordable Care Act.^26^ As we have shown here, Medicaid claims-based measures provide new opportunities to study PCU in this population. However, researchers using either source of data need to be aware of its potential benefits and limitations in order to interpret their results correctly.

## Conclusion

Claims-based estimates of PCU are moderately concordant with self-reported rates at the state level. However, rates measured through Medicaid claims compared to self-report diverge in some groups, suggesting they may measure different aspects of preconception care. Researchers using these two approaches to quantifying preconception care should understand the potential benefits and limitations of each.

## Data Availability

This is a secondary analysis of data received from the Centers for Medicare and Medicaid Services under an approved Data Use Agreement which governs all requests for data sharing.

## Tables and Figures

**Supplemental Table 1.** Logistic regressions of preconception care measures from 2012 PRAMS and Medicaid MAX on age, race/ethnicity and chronic conditions, estimated using data from 26 states available in PRAMS (odds ratios and 95% CI)

| Covariate | PRAMS 2012<sup>a</sup><br>(n = 6,960) | Medicaid MAX 2012<sup>b</sup><br>(n = 652,929) |  |  |
| --- | --- | --- | --- | --- |
|  | Preconception care | All Domains<sup>c</sup> | Contraceptive services | Related prev. health services |
| Age |  |  |  |  |
| ≤17 | 0.79<br>(0.53, 1.20) | 0.89‡<br>(0.86, 0.92) | 1.06†<br>(1.02, 1.10) | 0.53‡<br>(0.50, 0.55) |
| 18–19 | 0.75<br>(0.54, 1.03) | 1.16‡<br>(1.13, 1.18) | 1.24‡<br>(1.22, 1.27) | 0.88‡<br>(0.85, 0.90) |
| 20–24 | Ref. | Ref. | Ref. | Ref. |
| 25–29 | 1.17<br>(0.92, 1.48) | 0.90‡<br>(0.89, 0.91) | 0.80‡<br>(0.78, 0.81) | 1.13‡<br>(1.11, 1.15) |
| 30–34 | 0.98<br>(0.75, 1.29) | 0.76‡<br>(0.75, 0.77) | 0.56‡<br>(0.55, 0.58) | 1.06‡<br>(1.04, 1.08) |
| 35–39 | 1.01<br>(0.68, 1.49) | 0.65‡<br>(0.64, 0.67) | 0.40‡<br>(0.39, 0.42) | 0.98<br>(0.96, 1.01) |
| 40+ | 0.62<br>(0.33, 1.19) | 0.57‡<br>(0.55, 0.60) | 0.27‡<br>(0.25, 0.30) | 0.93*<br>(0.88, 0.98) |
| Race/ethnicity |  |  |  |  |
| White non-Hispanic | Ref. | Ref. | Ref. | Ref. |
| Black non-Hispanic | 1.99‡<br>(1.58, 2.50) | 1.50‡<br>(1.48, 1.52) | 1.30‡<br>(1.28, 1.32) | 1.36‡<br>(1.34, 1.38) |
| Hispanic | 2.01‡<br>(1.55, 2.61) | 0.84‡<br>(0.83, 0.85) | 0.87‡<br>(0.85, 0.89) | 0.89‡<br>(0.88, 0.91) |
| Asian/Pacific Islander | 3.08‡<br>(2.20, 4.33) | 0.79‡<br>(0.77, 0.82) | 0.49‡<br>(0.47, 0.52) | 1.00<br>(0.97, 1.04) |
| Other | 1.97‡<br>(1.41, 2.76) | 1.21‡<br>(1.17, 1.25) | 1.43‡<br>(1.37, 1.48) | 0.87‡<br>(0.83, 0.91) |
| Chronic Conditions |  |  |  |  |
| Diabetes | 1.87†<br>(1.19, 2.94) | 1.37‡<br>(1.31, 1.43) | 1.06<br>(1.00, 1.12) | 1.41‡<br>(1.35, 1.48) |
| Hypertension | 1.87†<br>(1.31, 2.67) | 1.25‡<br>(1.21, 1.30) | 1.10‡<br>(1.05, 1.16) | 1.28‡<br>(1.23, 1.33) |
| Depression | 1.08<br>(0.83, 1.40) | 1.82‡<br>(1.79, 1.86) | 1.61‡<br>(1.57, 1.65) | 1.64‡<br>(1.60, 1.68) |

\* p < 0.05; † p < 0.01; ‡ p < 0.001
<sup>a</sup> Includes only those who were Medicaid-eligible.
<sup>b</sup> Includes only those states for which PRAMS data were available.
<sup>c</sup> Includes all individuals with at least one claim during the year before conception in the following 7 domains: contraceptive services, pregnancy testing and counseling, natural family planning counseling (V26.41), basic infertility services, preconception health services, STD services, and related preventative health services.

**Supplemental Table 2.** Mixed-effects logistic regressions of preconception care measures from 2012 Medicaid MAX on age, race/ethnicity and chronic conditions, estimated using data from 46 states (odds ratios and 95% CI)^a^

| Covariate | Medicaid MAX 2012<sup>b</sup> (n = 1,452,034) |  |  |
| --- | --- | --- | --- |
|  | All Domains<sup>c</sup> | Contraceptive services | Related prev. health services |
| Age |  |  |  |
| ≤17 | 0.89‡<br>(0.87, 0.91) | 0.91‡<br>(0.88, 0.93) | 0.61‡<br>(0.59, 0.63) |
| 18–19 | 1.19‡<br>(1.17, 1.20) | 1.16‡<br>(1.14, 1.18) | 1.02<br>(1.00, 1.04) |
| 20–24 | Ref. | Ref. | Ref. |
| 25–29 | 0.85‡<br>(0.84, 0.85) | 0.79‡<br>(0.78, 0.80) | 1.05‡<br>(1.04, 1.06) |
| 30–34 | 0.69‡<br>(0.68, 0.70) | 0.57‡<br>(0.56, 0.57) | 0.95‡<br>(0.93, 0.96) |
| 35–39 | 0.58‡<br>(0.57, 0.59) | 0.41‡<br>(0.40, 0.42) | 0.87‡<br>(0.85, 0.89) |
| 40+ | 0.50‡<br>(0.49, 0.52) | 0.29‡<br>(0.27, 0.31) | 0.83‡<br>(0.79, 0.86) |
| Race/ethnicity |  |  |  |
| White non-Hispanic | Ref. | Ref. | Ref. |
| Black non-Hispanic | 1.60‡<br>(1.58, 1.61) | 1.50‡<br>(1.48, 1.52) | 1.38‡<br>(1.36, 1.40) |
| Hispanic | 0.70‡<br>(0.69, 0.70) | 0.77‡<br>(0.76, 0.78) | 0.69‡<br>(0.68, 0.70) |
| Asian/Pacific Islander | 0.67‡<br>(0.65, 0.68) | 0.53‡<br>(0.51, 0.55) | 0.79‡<br>(0.77, 0.81) |
| Other | 1.15‡<br>(1.12, 1.18) | 1.13‡<br>(1.10, 1.17) | 0.86‡<br>(0.83, 0.89) |
| Chronic Conditions |  |  |  |
| Diabetes | 1.41‡<br>(1.36, 1.45) | 1.12‡<br>(1.07, 1.17) | 1.47‡<br>(1.41, 1.52) |
| Hypertension | 1.27‡<br>(1.24, 1.31) | 1.14‡<br>(1.10, 1.19) | 1.31‡<br>(1.27, 1.36) |
| Depression | 1.74‡<br>(1.71, 1.77) | 1.56‡<br>(1.53, 1.59) | 1.60‡<br>(1.57, 1.63) |
| State-level random effect |  |  |  |
| SD | 0.42‡<br>(0.34, 0.52) | 0.48‡<br>(0.39, 0.59) | 0.47‡<br>(0.38, 0.57) |

\* p < 0.05; † p < 0.01; ‡ p < 0.001
<sup>a</sup> Models include state-level means of all covariates.
<sup>b</sup> Includes only those states for which PRAMS data were available.
<sup>c</sup> Includes all individuals with at least one claim during the year before conception in the following 7 domains: contraceptive services, pregnancy testing and counseling, natural family planning counseling (V26.41), basic infertility services, preconception health services, STD services, and related preventative health services.

## Acknowledgments

The authors gratefully acknowledge Ashley McHugh for her contributions to data acquisition and project management.

## Funding

This work was support by the Agency for Healthcare Research and Quality (grant number R03 HS27027). The content is solely the responsibility of the authors and does not necessarily represent the official views of the National Institutes of Health.

## Notes

### Competing Interest Statement

The authors have declared no competing interest.

### Funding Statement

This study was funded by the Agency for Healthcare Research and Quality (grant number R03 HS27027). The content is solely the responsibility of the authors and does not necessarily represent the official views of the National Institutes of Health.

### Author Declarations

IRB of the University of Chicago gave ethical approval for this work

